# Comorbid type 2 diabetes and chronic gastroduodenitis synergistically increase adverse clinical outcomes: implications for MRI-derived phenotype-tailored dietary strategies

**DOI:** 10.64898/2026.06.01.26354665

**Authors:** Yu-Ling Cui, Ying Yu, Guang-Bin Cui, Bo Hu

## Abstract

**Background:** Chronic gastritis and duodenitis (CGD) are highly prevalent among patients with type 2 diabetes (T2D). However, the prognostic impact of their comorbidity and the potential role of MRI-derived phenotype-tailored dietary strategies remain unclear.

**Methods:** This prospective cohort study included 453,768 UK Biobank participants. Primary endpoints were myocardial infarction, stroke, end-stage renal disease (ESRD), dementia, Parkinson’s disease, and all-cause mortality. Time-dependent multivariable Cox regression assessed outcome associations, while additive interaction analyses evaluated synergistic effects between T2D and CGD. Eight healthy dietary pattern scores were analyzed. Latent profile analysis classified MRI-derived body composition phenotypes based on fat distribution and organ volume.

**Results:** T2D and CGD were positively associated, and their comorbidity increased risks of cardiovascular events, ESRD, dementia, and all-cause mortality. Additive interaction analyses demonstrated synergistic effects on myocardial infarction and all-cause mortality. The comorbidity was further associated with aggravated lipid metabolic abnormalities and multiorgan atrophy. Higher adherence to the Healthful Plant-Based Diet Index (HPDI) and Dietary Approaches to Stop Hypertension (DASH) diets attenuated the excess mortality risk related to this synergy. Dietary associations varied across T2D, CGD, and comorbid populations, while MRI-based latent profiles modified diet-outcome relationships. A nomogram integrating demographic, dietary, and body composition data demonstrated reliable long-term predictive performance for myocardial infarction, stroke, and all-cause mortality.

**Conclusions:** Comorbid T2D and CGD substantially increase adverse clinical risks and exhibit synergistic effects on myocardial infarction and all-cause mortality. These findings support routine CGD screening in T2D care and provide population-based evidence for MRI-derived phenotype-tailored dietary strategies.

## 1. Introduction

Type 2 diabetes (T2D) is a major global health burden, accounting for a substantial proportion of preventable deaths worldwide (1). Comorbid conditions substantially influence long-term clinical outcomes in individuals with T2D (2). Nevertheless, current clinical research continues to emphasize the management of classic cardiovascular, renal, and ophthalmic complications, often overlooking other highly prevalent yet underrecognized comorbidities. Among these is chronic gastritis and duodenitis (CGD) (3), which exhibits a significantly higher prevalence in patients with T2D compared with the general population (4). However, the impact of comorbid T2D and CGD on long-term prognosis remains unclear, and targeted intervention strategies for this comorbidity are still lacking.

Chronic hyperglycemia compromises gastrointestinal mucosal integrity and increases susceptibility to *Helicobacter pylori* (*H. pylori*) infection (5–7), whereas CGD-related chronic low-grade inflammation and gut microbial dysbiosis in turn exacerbate insulin resistance and impair glycemic control (8–11). Although this bidirectional relationship is well documented, few clinical studies have quantified their additive synergistic interaction (12), namely, whether comorbidity elevates adverse event risk beyond the combined effects of each condition individually. If confirmed, this evidence would help optimize clinical care for patients with T2D by integrating routine CGD screening and, more importantly, guide targeted clinical interventions.

Healthy dietary patterns represent a cornerstone interventional strategy capable of simultaneously improving metabolic regulation and mitigating intestinal inflammation (13–15). Accordingly, identifying dietary patterns that optimally enhance clinical outcomes in patients with comorbid T2D and CGD holds profound clinical relevance (16). However, prior investigations failed to meet the rigorous demands of targeted dietary interventions, largely due to their inadequate accounting for holistic systemic nutritional and metabolic status (17–20). Whole-body adipose tissue distribution and abdominal organ volume are tightly linked to systemic nutritional and metabolic status (21, 22), and thus represent critical, underutilized phenotypes for patient stratification. While recent studies have applied MRI-derived features of whole-body adipose tissue distribution in conjunction with latent profile analysis (LPA) to stratify patient populations, these analyses have omitted organ volume metrics (23, 24). Consequently, integrating MRI features of both whole-body adipose tissue distribution and organ volume for LPA would enable targeted risk stratification and dietary interventions, thereby carrying strong clinical utility and translational potential.

To address these critical evidence gaps, we performed a prospective cohort analysis using data from the UK Biobank. We hypothesized that: (1) the coexistence of T2D and CGD is associated with an elevated risk of six prespecified major adverse clinical outcomes, and exerts a significant additive synergistic interaction effect on these outcomes; (2) healthy dietary patterns are associated with improved prognosis and attenuate this synergistic comorbidity-related risk; (3) body composition subtypes identified via MRI enable the identification of subgroups responsive to dietary interventions; and (4) a nomogram integrating demographic and clinical data, dietary quality scores, and body composition subtypes exhibits robust and visualizable prognostic performance. Validation of these hypotheses will provide high-level evidence to guide risk stratification and targeted dietary strategies for patients with T2D, CGD, and the comorbidity.

## 2. Methods

### 2.1 Study design

This study utilized data from the UK Biobank (application no. 627927), a large-scale, ongoing prospective cohort study encompassing over 500,000 participants in the United Kingdom (25). Written informed consent was obtained from all participants, and ethical approval was granted by the North West Multicenter Research Ethics Committee (21/NW/0157).

All variables were collected through standardized questionnaires and physical measurements. The demographic and lifestyle-related variables included age, sex, body mass index (BMI), Townsend deprivation index, home location, smoking frequency, alcohol consumption frequency, and sleep duration.

### 2.2 Participant inclusion and exclusion

The participant inclusion and exclusion flowchart is shown in **Figure 1**. A total of 502,128 individuals were initially recruited from the UK Biobank, and stepwise exclusion criteria were applied to establish the final analytical cohort. First, 39,684 participants were excluded due to conditions that may confound the exposure-outcome association, including type 1 diabetes, malnutrition-related diabetes, gestational diabetes, acute gastritis, Crohn’s disease, ulcerative colitis, schizophrenia, malignancy, heart failure, respiratory failure, multiple sclerosis, brain trauma and HIV infection. An additional 8,676 individuals were excluded if they had a baseline history of primary outcomes, including myocardial infarction, stroke, end-stage renal disease (ESRD), dementia, and Parkinson’s disease. After exclusions, 453,768 eligible participants were retained and divided into four cohorts: (1) Control cohort (no T2D or CGD), (2) T2D-only cohort, (3) CGD-only cohort, and (4) comorbidity (COM) cohort (both T2D and CGD). The median follow-up period was 16.12 years. Analyses of dietary data included 193,686 participants with complete dietary assessments, while analyses of MRI data included 15,178 participants with valid MRI scans.

**Fig. 1.**
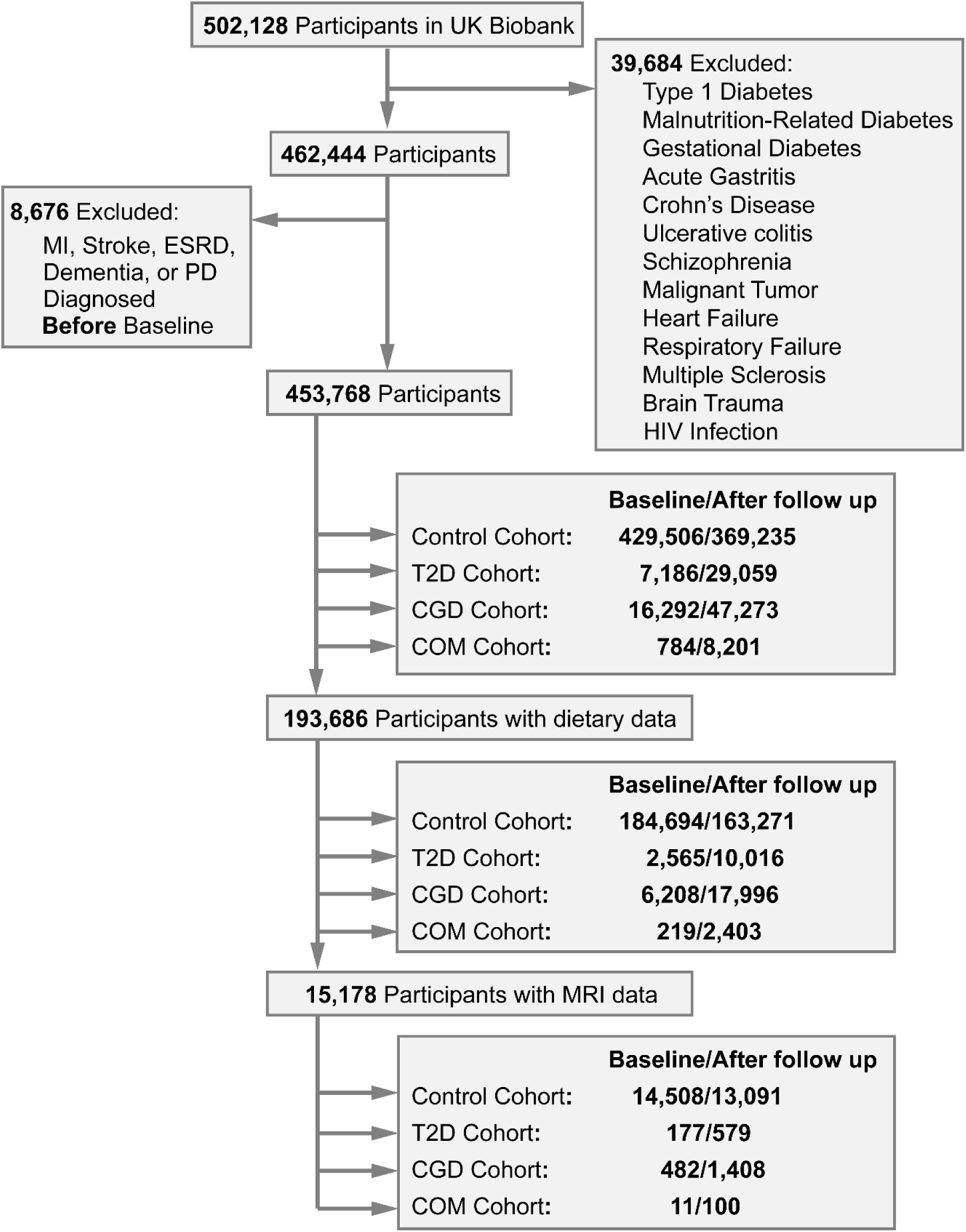
Flowchart of participant recruitment and subgrouping. The number of participants in each group is presented both at baseline and upon completion of follow-up, corresponding to the sample used for the time-dependent Cox regression model.

### 2.3 Diagnosis of T2D and CGD

The UK Biobank derives disease onset dates from multiple linked data sources, including hospital episode statistics, primary care records, and the death register. T2D was diagnosed using any of the following criteria: (1) a recorded diagnosis based on the International Classification of Diseases, Tenth Revision (ICD–10) code E11 (non-insulin-dependent diabetes mellitus); (2) a baseline fasting blood glucose level ≥ 11.1 mmol/L; or (3) a baseline glycated hemoglobin (HbA1c) level ≥ 6.5% (48 mmol/mol). CGD was diagnosed based on ICD–10 code K29 (gastritis and duodenitis), after excluding codes K29.0 (acute hemorrhagic gastritis) and K29.1 (other acute gastritis).

### 2.4 Primary outcomes

The primary outcomes consisted of six common T2D-related clinical adverse events: myocardial infarction, stroke, ESRD, dementia, Parkinson’s disease, and all–cause mortality. All outcomes were derived from UK Biobank health records, in which validated algorithms were applied to integrate self–reported conditions, hospital admission data, and death registry information for standardized case classification.

### 2.5 Dietary patterns

Dietary intake was assessed using the validated Oxford WebQ 24–hour recall instrument in the UK Biobank cohort. Based on average food and nutrient intakes, eight well-established dietary pattern scores were calculated (see **Supplementary Note 1** for detailed introduction):

*The anti–Empirical Dietary Inflammatory Index (AEDII)* (*26, 27*): a reversed pro–inflammatory score reflecting anti–inflammatory diet quality (detailed scoring criteria are provided in **Table S1)**.

*The Alternative Healthy Eating Index 2010 (AHEI–2010)* (*26, 28*): a measure of overall diet quality that encourages health–promoting foods and limits pro–inflammatory or adverse dietary components (**Table S2**).

*The Dietary Approaches to Stop Hypertension (DASH)* (*29, 30*): assessing adherence to a dietary pattern associated with lower risks of hypertension, cardiovascular disease, and related chronic conditions (**Table S3**).

*Two versions of the EAT-Lancet diet index* (*31*): developed to evaluate adherence to a healthy and sustainable dietary pattern supporting both human and environmental health. These include the Knuppel version (ELK, **Table S4**), which uses binary scoring, and the Stubbendorff version (ELS, **Table S5**), which uses an ordinal scale.

*The Mediterranean Diet Adherence Screener (MEDAS)* (*26*): characterized by a high intake of vegetables and fruits, regular consumption of fish and seafood, and moderate red wine intake (detailed scoring criteria are provided in **Ref. 26**).

*The Mediterranean-DASH Intervention for Neurodegenerative Delay diet (MIND)* (*26*): a neuroprotective dietary index combining principles from the Mediterranean and DASH diets, focused on slowing neurodegeneration (detailed scoring criteria are provided in **Ref. 26**).

*The Healthful Plant–Based Diet Index (HPDI)* (*26, 32, 33*): a plant-based score emphasizing the consumption of healthful plant foods (**Table S6**).

### 2.6 Imaging data

This analysis incorporated the eight standard abdominal fat-related MRI measures established in prior research (23): hepatic proton density fat fraction (PDFF), pancreatic PDFF, abdominal fat ratio, muscle fat infiltration, weight to muscle ratio, abdominal subcutaneous adipose tissue (ASAT) volume, visceral adipose tissue (VAT) volume, and pericardial fat area. Due to the significant impact of organ volume on metabolism, we expanded the body composition profile by including five organ volume metrics: liver volume, kidney volume, lung volume, spleen volume, and pancreatic volume. To minimize analytic bias, participants with extreme outlier values (≤ -5 standard deviations [SD] or ≥ 5 SD) on any MRI measure were excluded (23). Detailed protocols for UK Biobank MRI acquisition, fat quantification, and organ volume estimation have been previously published (34).

All MRI-based measurements of adipose tissue and visceral organs were preprocessed to derive quantitative indices independent of general adiposity and overall body size. Consistent with established analytical protocols in prior literature (23), regional fat-related metrics were residualized for body mass index (BMI). Since height and weight better reflect variations in organ volume compared with BMI, organ volume parameters were adjusted for height and weight, and both were processed via sex-stratified linear regression models.

The resultant residual values were subsequently standardized into sex-specific z-scores calculated as (x−mean)/SD, yielding fully adjusted indicators for downstream subgroup classification. Collinearity assessment was systematically performed across all included variables, and any factor with a variance inflation factor (VIF) exceeding 5 was pre-specified for exclusion. Notably, no variables were removed in the present analysis, as all VIF values remained below the threshold of 5.

### 2.7 Latent profile analysis of MRI-derived phenotypes

Latent profile analysis (LPA) was conducted using the transformed, non–collinear MRI fat and organ volume quantification variables using the R package tidyLPA 2.0.2. Model selection followed three criteria: (a) lower Bayesian and Akaike Information Criteria, indicating a better trade–off between fit and parsimony; (b) a local maximum in entropy (scale 0–1); and (c) no profile containing fewer than 5% of the total sample, to avoid overfitting.

### 2.8 Statistical analysis

#### 2.8.1 Descriptive statistics and group comparisons

Baseline demographic and clinical characteristics were summarized as frequencies and percentages for categorical variables and as means ± SDs for continuous variables. Differences in baseline characteristics among cohorts were evaluated using analysis of variance (ANOVA) for continuous variables and Chi-square tests for categorical variables. Between-profile disparities in dietary pattern scores were assessed via analysis of covariance (ANCOVA), with age, sex, BMI, Townsend deprivation index, smoking frequency, alcohol intake frequency, and sleep duration included as confounding covariates. Pairwise between-group effect sizes were quantified using Cohen’s *d*, with all calculations referenced to benchmark profile in the primary analyses.

#### 2.8.2 Survival analysis and multivariable Cox regression

The association between T2D and CGD was initially quantified using odds ratios (ORs). The cumulative incidence of primary outcomes across the cohorts was estimated via Kaplan-Meier survival analysis. To eliminate immortal time bias (35), the time-dependent Cox model was applied to examine the associations of T2D, CGD and their comorbidity with the risks of primary outcomes, with adjustment for age, sex, BMI and other baseline covariates. Proportional hazards were verified with Schoenfeld residuals (36). Results are presented as hazard ratios (HRs) with 95% confidence intervals (CIs). Statistical significance was defined as a two-sided P < 0.05, and the false discovery rate (FDR) was applied to correct for multiple comparisons.

#### 2.8.3 Assessment of synergistic and interaction effects

To evaluate whether the combined effect of T2D and CGD on the risk of primary outcomes exceeded the sum of their individual effects, we estimated three widely adopted additive-scale interaction metrics (see **Supplementary Note 2** for detailed introduction), including the relative excess risk due to interaction (RERI), the attributable proportion due to interaction (AP), and the synergy index (SI) (12). Bootstrap resampling with 5,000 iterations was employed to derive 95% CIs for these indices.

#### 2.8.4 Development and validation of the predictive nomogram

Time-dependent multivariable Cox regression models were built to predict primary outcomes using demographic, clinical, dietary and latent profile analysis (LPA) data. To maintain model parsimony and reduce overfitting, stepwise variable selection was conducted based on the Akaike Information Criterion (AIC), retaining only informative and statistically significant prognostic factors. A visual nomogram was then developed from the final optimized model to enable individualized prediction of endpoint risks.

The model’s predictive accuracy and discriminative ability were assessed using receiver operating characteristic (ROC) curves, precision-recall (PR) curves, calibration curves and decision curve analysis (DCA). The area under the ROC curve (AUC-ROC) and area under the PR curve (AUC-PR) were calculated accordingly.

## 3. Results

### 3.1 Baseline Characteristics of Study Participants

**Table 1** presents baseline demographic, clinical, dietary and MRI profiles of all enrolled participants stratified by cohort. The COM cohort exhibited significantly older age (60.89 ± 6.45), greater BMI (32.14 ± 6.02), higher Townsend deprivation index (60.89 ± 6.45) and prolonged sleep duration (7.29 ± 1.57) than the remaining three cohorts (all FDR-Corrected *P* < 0.001). Across all adverse endpoints, namely myocardial infarction, stroke, ESRD, dementia, Parkinson’s disease and all-cause mortality, event rates were ranked highest in the COM cohort, followed by the T2D cohort, the CGD cohort and the Control cohort. A significant positive correlation was identified between T2D and CGD (OR = 2.10, 95% CI 1.97-2.24), suggesting that the presence of T2D was associated with 2.1-fold increased odds of CGD occurrence.

**Table 1.**
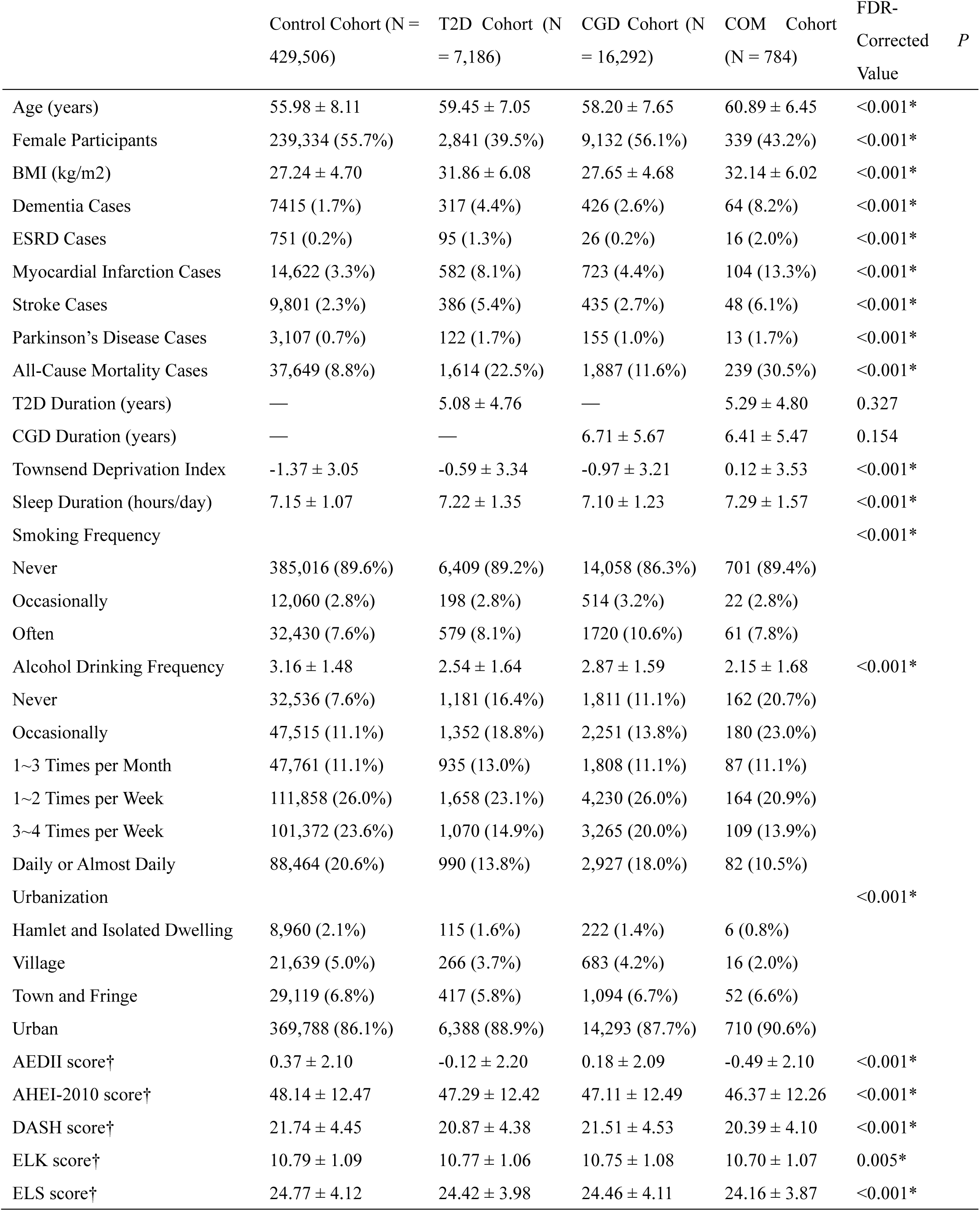
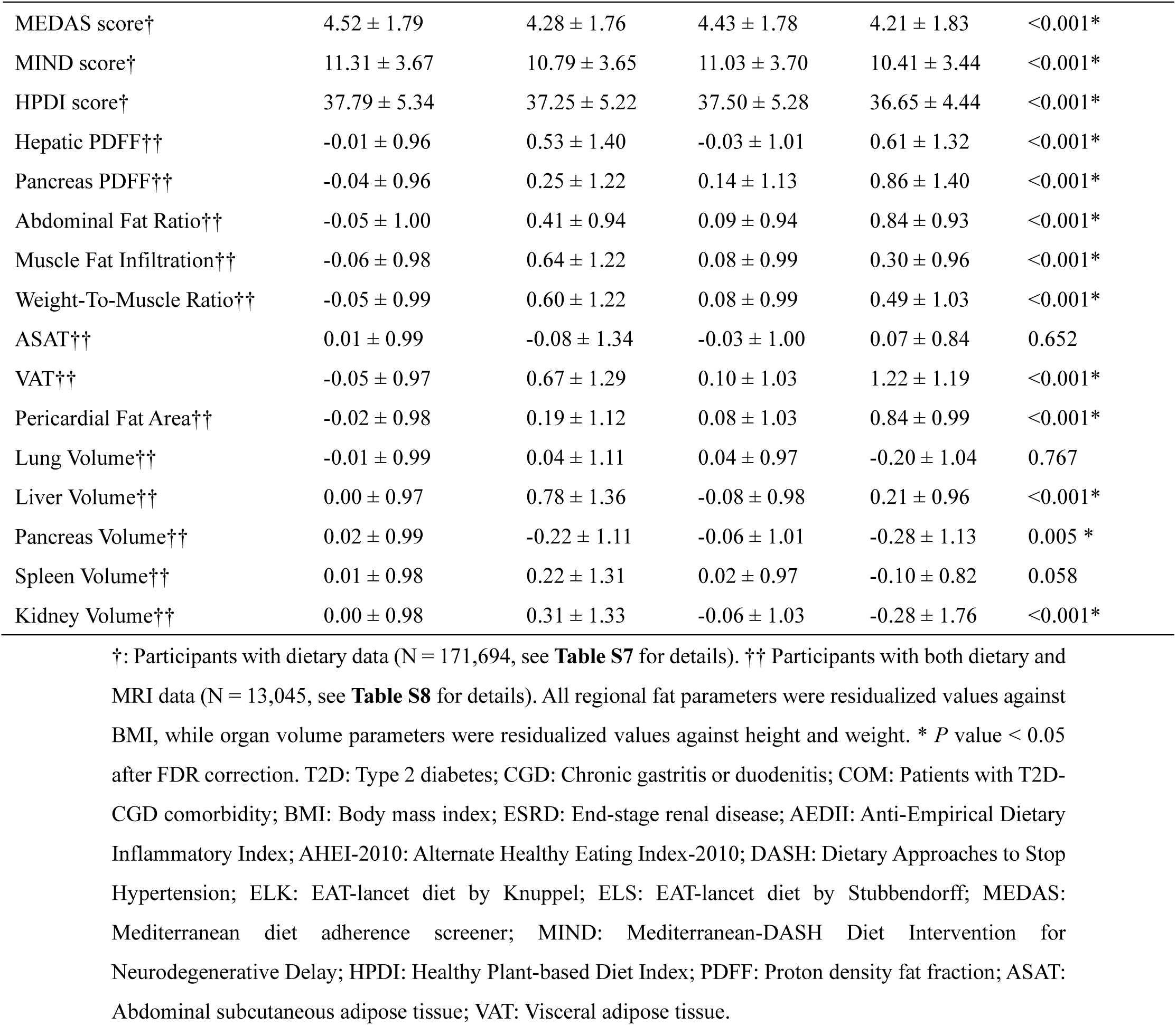
Demographic, clinical, and dietary characteristics.

Healthy dietary patterns revealed that COM cohort had the lowest adherence to healthy dietary patterns relative to the other cohorts, with values of −0.49 ± 2.10 for AEDII, 46.37 ± 12.26 for AHEI-2010, 20.39 ± 4.10 for DASH, 10.70 ± 1.07 for ELK, 24.16 ± 3.87 for ELS, 4.21 ± 1.83 for MEDAS, 10.41 ± 3.44 for MIND, and 36.65 ± 4.44 for HPDI, respectively (all FDR-Corrected *P* < 0.01).

Quantitative MRI analyses revealed that the COM cohort exhibited the highest hepatic (0.61 ± 1.32), pancreatic PDFF (0.86 ± 1.40), abdominal fat ratio (0.84 ± 0.93), VAT (1.22 ± 1.19) and pericardial fat area (0.84 ± 0.99) (all FDR-Corrected *P* < 0.001). Conversely, kidney volumes (−0.28 ± 1.76) and pancreatic volume (−0.28 ± 1.13) were lowest in this cohort (all FDR-Corrected *P* < 0.01). These observations imply that comorbid T2D and CGD aggravates systemic lipid metabolic dysfunction and atrophy of kidney and pancreas.

### 3.2 Association between T2D-CGD comorbidity and primary outcomes

The Kaplan-Meier survival curves in **Fig. 2** indicate that the COM cohort exhibited the highest cumulative incidence of myocardial infarction, stroke, ESRD, dementia, Parkinson’s disease, and the lowest overall survival (log-rank test *P* < 0.0001). Based on the time-dependent multivariable Cox regression analysis that adjusted all covariates (**Table 2**), comorbid T2D and CGD were significantly associated with an elevated risk of myocardial infarction (HR = 2.06, 95% CI 1.87-2.27), stroke (HR = 1.79, 95% CI 1.58-2.03), ESRD (HR = 4.98, 95% CI 3.72-6.67), dementia (HR = 3.13, 95% CI 2.78-3.53), Parkinson’s disease (HR = 1.81, 95% CI 1.44-2.26), and all-cause mortality (HR = 3.82, 95% CI 3.63-4.02). Results from additional models adjusting for different sets of covariates were consistent with the primary findings (**Table S9-S10**).

**Fig. 2.**
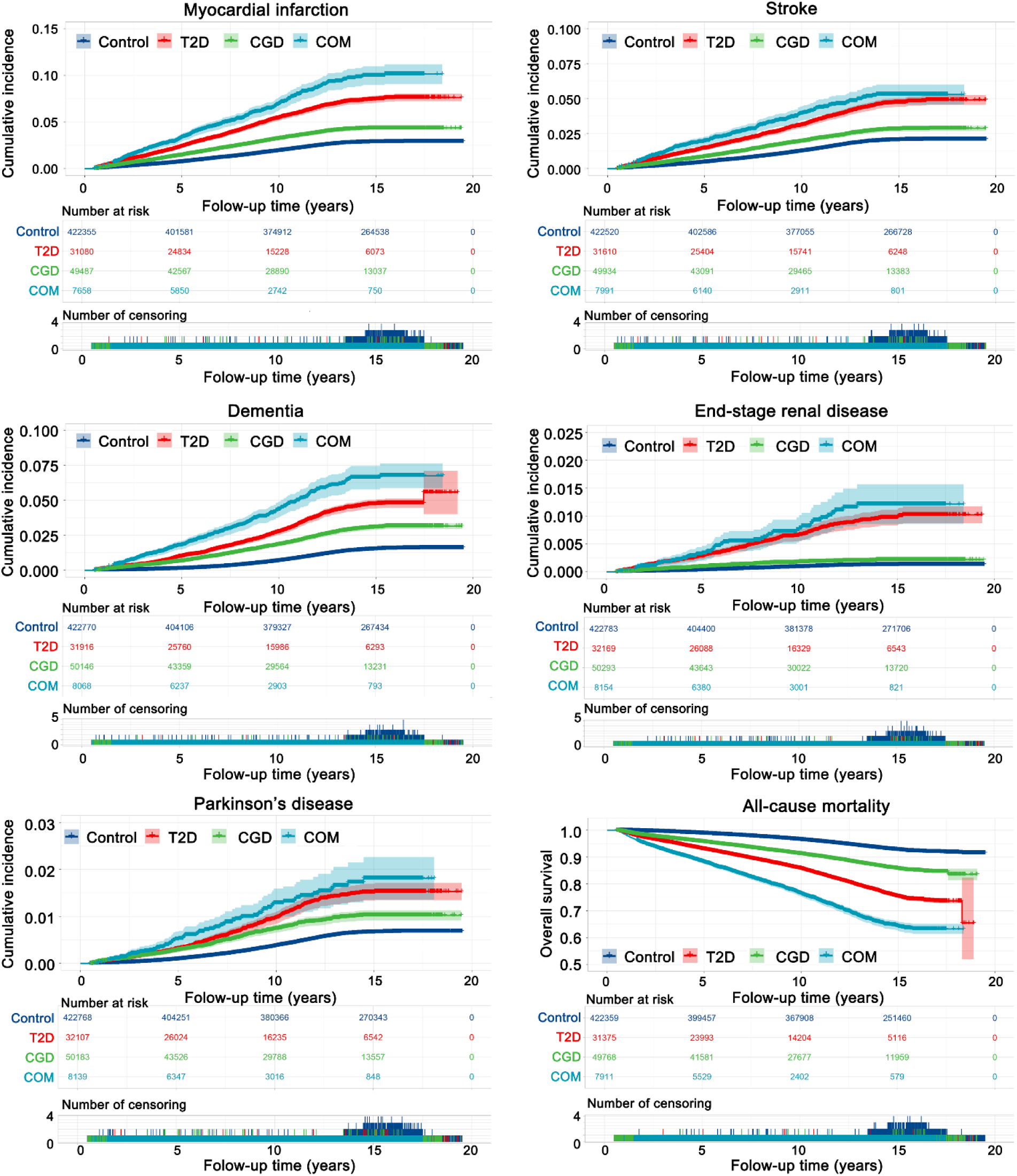
Comparison of primary outcomes by patient cohort. Kaplan-Meier curves show the primary outcomes for patients with type 2 diabetes and comorbidity (COM cohort), type 2 diabetes alone (T2D cohort), chronic generalized disease alone (CGD cohort), and control patients with neither condition (Control cohort).

**Table 2.** Hazard ratios (HRs) from Cox regression models adjusted for CGD/T2D status and duration, age, sex, BMI, sleep duration, smoke and drink frequency and Townsend deprivation index.

|  | T2D-CGD<br>Comorbidity | T2D<br>Alone | CGD<br>Alone | Age | Male Sex | BMI | Townsend<br>Deprivation<br>Index | Urbanization<br>Level | Sleep<br>Duration | Smoke<br>Frequency | Drink<br>Frequency |
| --- | --- | --- | --- | --- | --- | --- | --- | --- | --- | --- | --- |
| Myocardial<br>Infarction | 2.06 [1.87,<br>2.27] | 1.59<br>[1.51,<br>1.68] | 1.32 [1.25,<br>1.39] | 1.06 [1.06,<br>1.07] | 2.53<br>[2.45,<br>2.62] | 1.04 [1.03,<br>1.04] | 1.02 [1.01,<br>1.02] | 0.99 [0.96,<br>1.02] | 0.98 [0.96,<br>0.99] | 1.45 [1.42,<br>1.49] | 0.92 [0.91,<br>0.93] |
| Stroke | 1.79 [1.58,<br>2.03] | 1.57<br>[1.46,<br>1.68] | 1.14 [1.07,<br>1.22] | 1.09 [1.09,<br>1.10] | 1.44<br>[1.39,<br>1.50] | 1.02 [1.01,<br>1.02] | 1.03 [1.02,<br>1.04] | 1.01 [0.98,<br>1.04] | 0.99 [0.98,<br>1.01] | 1.36 [1.32,<br>1.40] | 0.98 [0.96,<br>0.99] |
| ESRD | 4.98 [3.72,<br>6.67] | 4.53<br>[3.79,<br>5.41] | 1.51 [1.20,<br>1.91] | 1.04 [1.03,<br>1.05] | 1.96<br>[1.70,<br>2.26] | 1.04 [1.03,<br>1.05] | 1.05 [1.02,<br>1.07] | 1.04 [0.92,<br>1.18] | 1.06 [1.00,<br>1.12] | 1.13 [1.01,<br>1.26] | 0.86 [0.83,<br>0.90] |
| Dementia | 3.13 [2.78,<br>3.53] | 2.18<br>[2.03,<br>2.35] | 1.60 [1.49,<br>1.70] | 1.20 [1.20,<br>1.21] | 1.17<br>[1.12,<br>1.22] | 0.99 [0.99,<br>1.00] | 1.03 [1.03,<br>1.04] | 1.07 [1.03,<br>1.11] | 1.00 [0.98,<br>1.02] | 1.13 [1.09,<br>1.18] | 0.93 [0.92,<br>0.95] |
| Parkinson's<br>Disease | 1.81 [1.44,<br>2.26] | 1.56<br>[1.38,<br>1.76] | 1.33 [1.20,<br>1.49] | 1.13 [1.13,<br>1.14] | 1.85<br>[1.72,<br>1.99] | 1.00 [0.99,<br>1.00] | 1.01 [1.00,<br>1.02] | 0.97 [0.92,<br>1.02] | 1.04 [1.01,<br>1.07] | 0.84 [0.78,<br>0.91] | 0.94 [0.92,<br>0.96] |
| All-cause<br>Mortality | 3.82 [3.63,<br>4.02] | 2.44<br>[2.36,<br>2.51] | 1.71 [1.66,<br>1.76] | 1.11 [1.11,<br>1.11] | 1.55<br>[1.52,<br>1.58] | 1.02 [1.02,<br>1.02] | 1.04 [1.04,<br>1.04] | 1.01 [1.00,<br>1.03] | 1.01 [1.00,<br>1.02] | 1.62 [1.60,<br>1.64] | 0.98 [0.97,<br>0.98] |

T2D: Type 2 diabetes; CGD: Chronic gastritis or duodenitis; ESRD: End-stage renal disease; BMI: Body mass index; TDI: Townsend deprivation index.

### 3.3 The synergistic effect between T2D and CGD on primary outcomes

The synergistic effect of T2D and CGD on the risk of myocardial infarction (RERI = 1.70, 95% CI 1.13-2.40; AP = 0.35, 95% CI 0.26-0.44; SI = 1.81, 95% CI 1.52-2.17) and all-cause mortality (RERI = 1.23, 95% CI 0.76-1.76; AP = 0.34, 95% CI 0.24-0.43; SI = 1.81, 95% CI 1.52-2.17) surpassed the sum of their individual effects. However, the synergistic effect of T2D and CGD on the risk of stroke, ESRD, dementia, and Parkinson’s disease did not exceed the sum of their individual effects (**Table S11**).

Using the observed number of events and the distribution of comorbid status, we estimated the minimum detectable HR with 80% power at a two-sided *α* of 0.05. For stroke (N = 10,317), ESRD (N = 858), dementia (N = 3,358), and Parkinson’s disease (N = 180), the detectable threshold HRs were 1.10, 1.38, 1.11, and 1.16 when comparing comorbid T2D-CGD with isolated T2D. The observed HRs for synergistic effects were 1.14 for stroke, 1.10 for ESRD, 1.44 for dementia, and 1.17 for Parkinson’s disease. Consequently, the risk for ESRD fell well below the detectable threshold. This suggests that the present study lacked sufficient statistical power to identify small-to-moderate synergistic effects, attributable to the limited number of outcome events.

### 3.4 The association between healthy dietary patterns and primary outcomes

The association between all healthy dietary patterns and primary outcomes is shown in **Fig. 3**. In the T2D cohort (**Table S12**), MEDAS diet was most protective against myocardial infarction (HR = 0.93, 95% CI 0.88-0.97, FDR-corrected *P* = 0.010), dementia (HR = 0.91, 95% CI 0.85-0.97, FDR-corrected *P* = 0.013), and all-cause mortality (HR = 0.94, 95% CI 0.90-0.98, FDR-corrected *P* = 0.027).

**Fig. 3.**
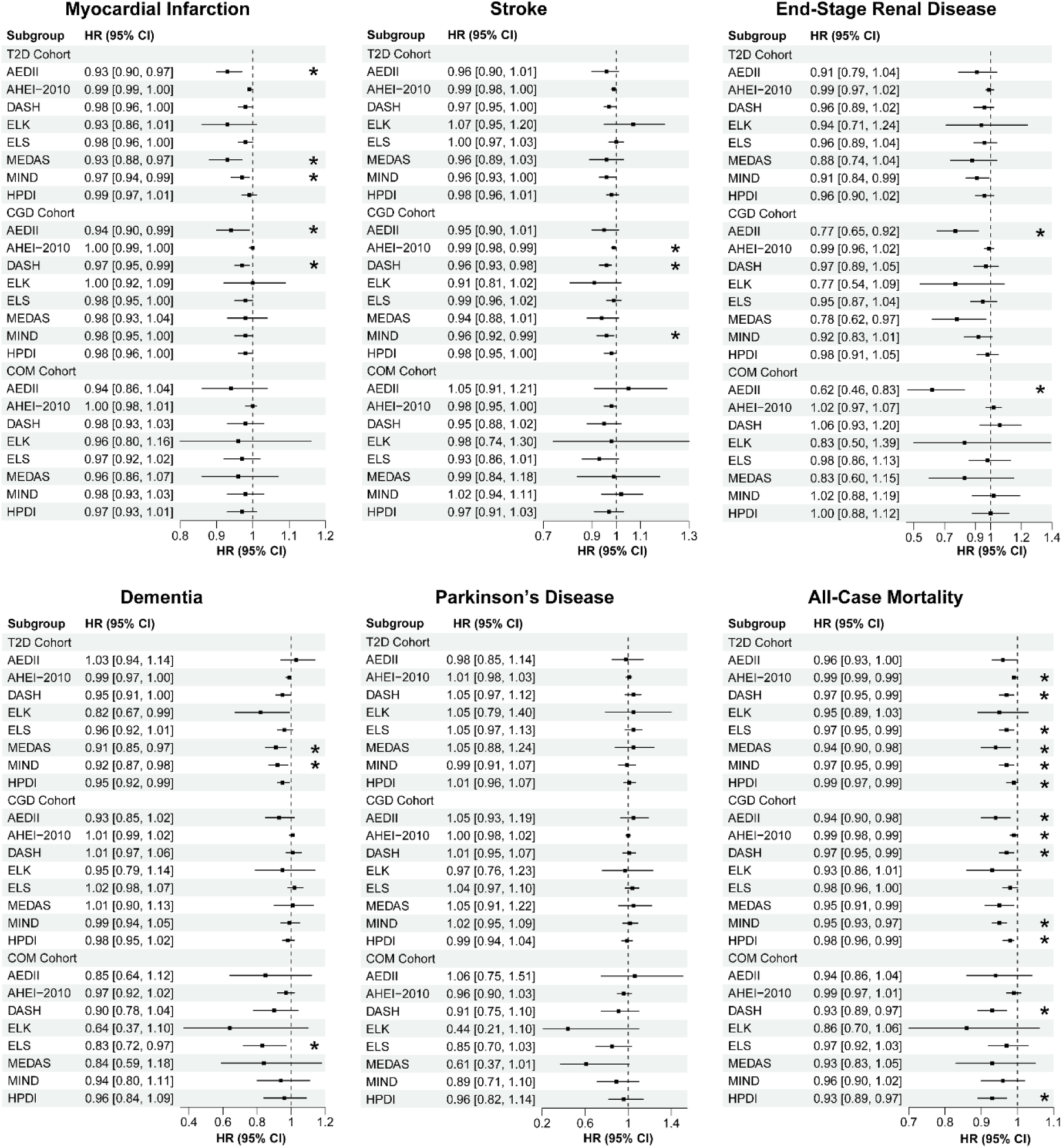
The association between healthy dietary patterns and primary outcomes by patient cohort. Forest plots show the associations (hazard ratios) between primary outcomes and dietary patterns for patients with type 2 diabetes and comorbidity (COM cohort), type 2 diabetes alone (T2D cohort), chronic generalized disease alone (CGD cohort), and control patients with neither condition (Control cohort). *: FDR-corrected *P* value <0.05.

In the CGD cohort (**Table S13**), AEDII diet was most protective against myocardial infarction (HR = 0.94, 95% CI 0.90-0.99, FDR-corrected *P* = 0.029), ESRD (HR = 0.77, 95% CI 0.65-0.92, FDR-corrected *P* =0.016), and all-cause mortality (HR = 0.94, 95% CI 0.9-0.98, FDR-corrected *P* = 0.008). DASH diet was most protective against stroke (HR =0.96, 95% CI 0.93-0.98, FDR-corrected *P* = 0.010).

In the COM cohort (**Table S14**), AEDII diet was most protective against ESRD (HR = 0.62, 95% CI 0.46-0.83, FDR-corrected *P* = 0.008). ELS diet was most protective against dementia (HR = 0.83, 95% CI 0.72-0.97, FDR-corrected *P* = 0.039). DASH diet was most protective against all-cause mortality (HR = 0.93, 95% CI 0.89-0.97, FDR-corrected *P* = 0.025).

In the Control cohort (**Table S15**), MEDAS diet showed protective effects against all six outcome events. Additionally, the protective effects of these dietary patterns showed no significant differences between female (**Table S16**) and male cohorts (**Table S17**).

### 3.5 The modulatory role of dietary pattern on the synergistic effect between T2D and CGD

The above experiments demonstrated that the synergistic effect of T2D and CGD on all-cause mortality exceeded the sum of their individual effects. Additionally, higher adherence to both HPDI and DASH dietary patterns was associated with a significantly lower risk of all-cause mortality (**Fig. 3**), suggesting that these dietary patterns may modulate the synergistic effect of T2D and CGD on mortality.

To test this hypothesis, a generalized linear model was applied, and three-way interaction analyses were conducted (**Fig. 4B and 4C**) (see **Supplementary Note 3** for details). The interaction between HPDI diet score, T2D, and CGD was statistically significant (*P* = 0.009). Consistently, the interaction between DASH diet score, T2D, and CGD also reached statistical significance (*P* = 0.014). The results show that HPDI and DASH diets could attenuate the synergistic effect between T2D and CGD.

**Fig. 4.**
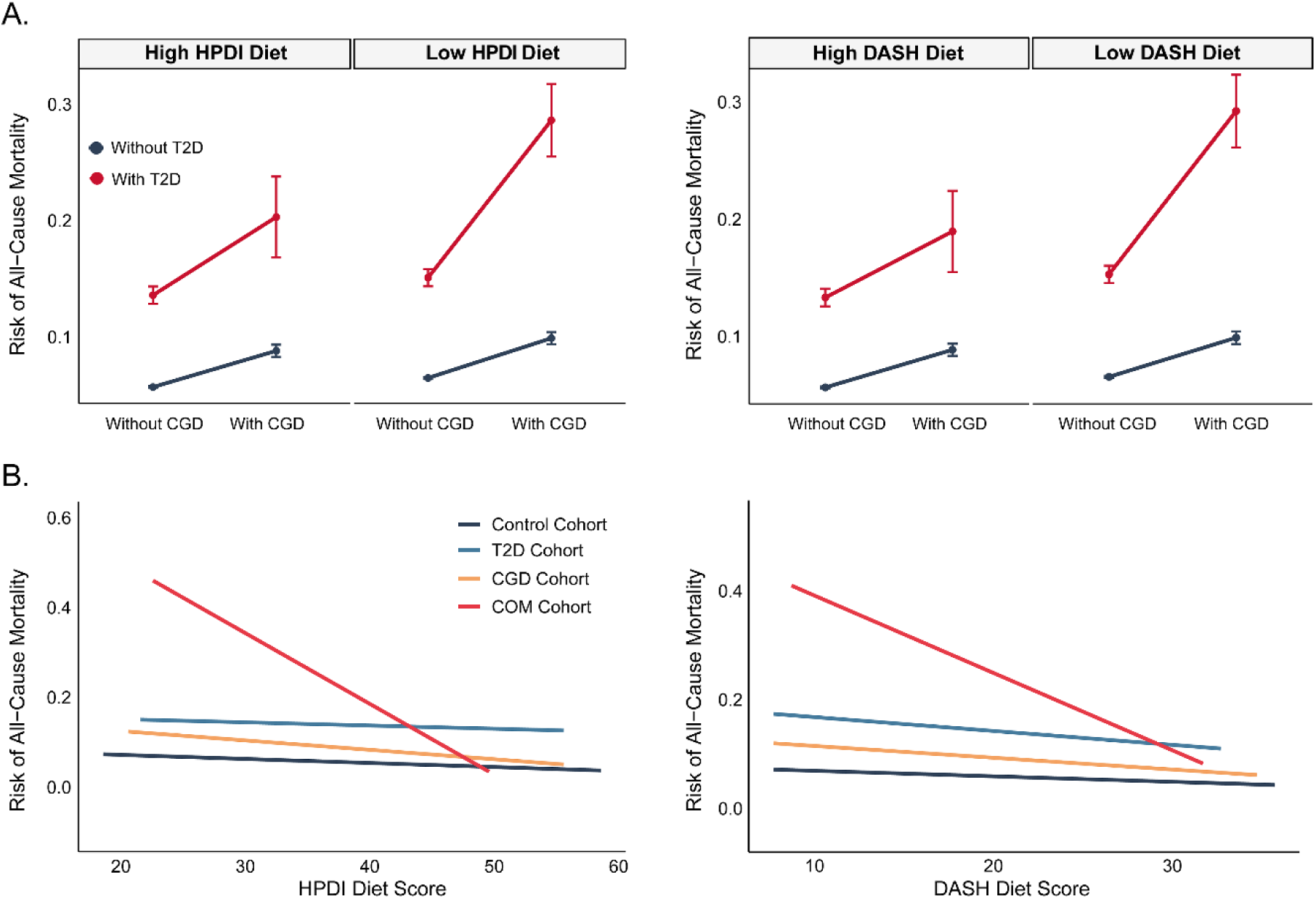
Three-way interactions between diet scores, T2D and CGD. (A) HPDI and (B) DASH diets weakened the synergistic effect of T2D and CGD on the outcome of all-cause mortality.

### 3.6 Identification of six latent profiles using fat distribution and organ volume MRI features

LPA incorporating fat distribution and organ volume MRI parameters identified six fat distribution patterns (**Fig. 5A**, detailed parameters for subtype recognition are displayed in **Table S18**). Significant inter-profile differences were observed in age, BMI, cardiometabolic disease prevalence, lifestyle, Townsend deprivation index and MRI characteristics in both sexes (**Table S19**). As shown in **Table 3**, Profile 4 (Lean) was associated with the highest scores on the DASH, AHEI-2010, ELS, and HPDI diets, whereas Profile 1 (Hepatocyte-Predominant) and Profile 6 (Balanced High Adiposity) were associated with the lowest dietary quality scores.

**Fig. 5.**
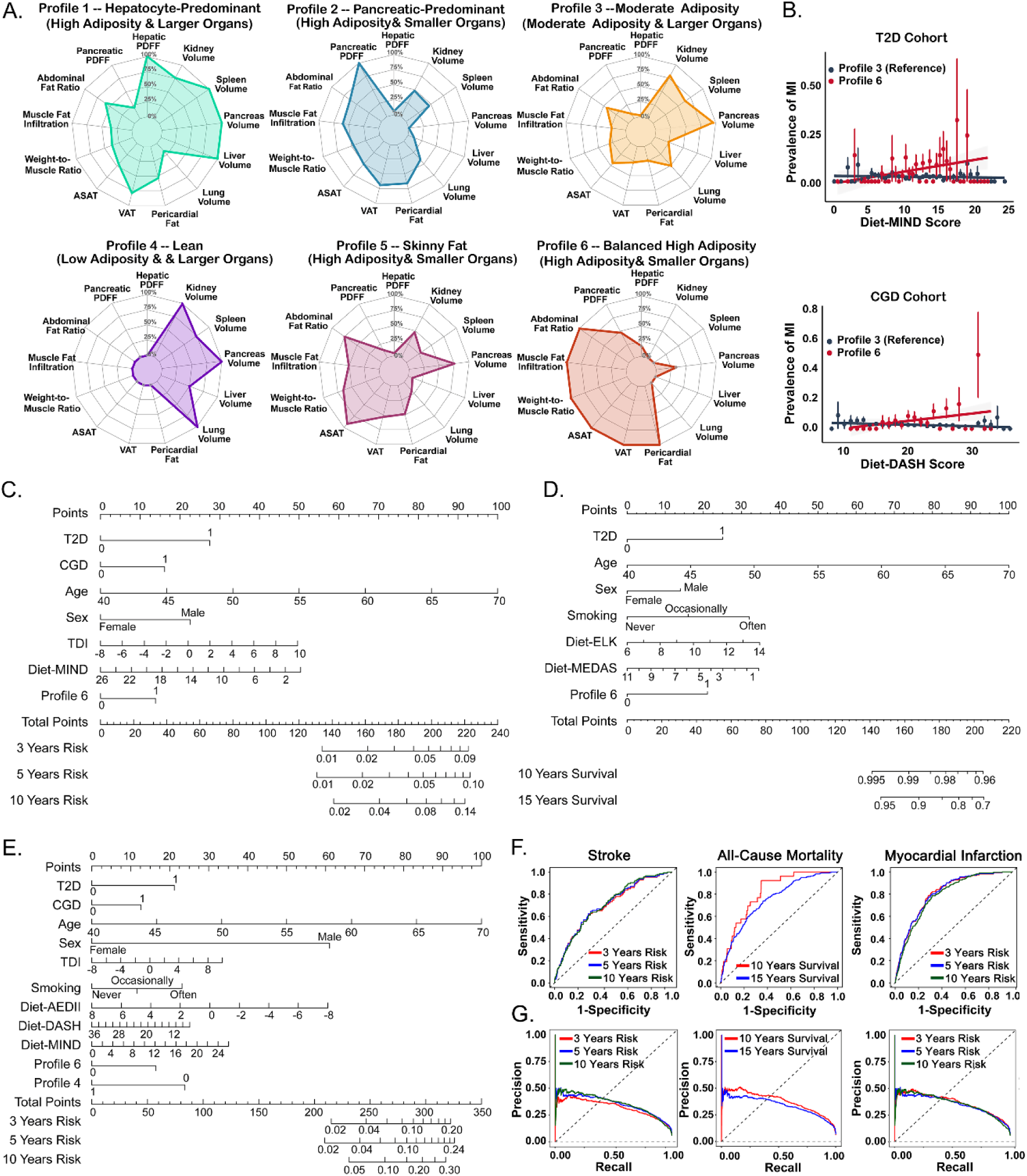
LPA classification, interaction effects and predictive nomograms. (A) Six adiposity profiles were defined using MRI features. Inter-profile differences in clinical, lifestyle and dietary indicators were observed in **Table 3**. (B) LPA profiles modified the association between dietary patterns and myocardial infarction risk. Significant interactions between diet and Profile 6 were identified in T2D and CGD cohorts after FDR correction. (C-E) Time-dependent Cox regression-based nomograms for stroke, all-cause mortality and myocardial infarction. (F-G) Model performance evaluated via ROC and PRC AUCs.

**Table 3.**
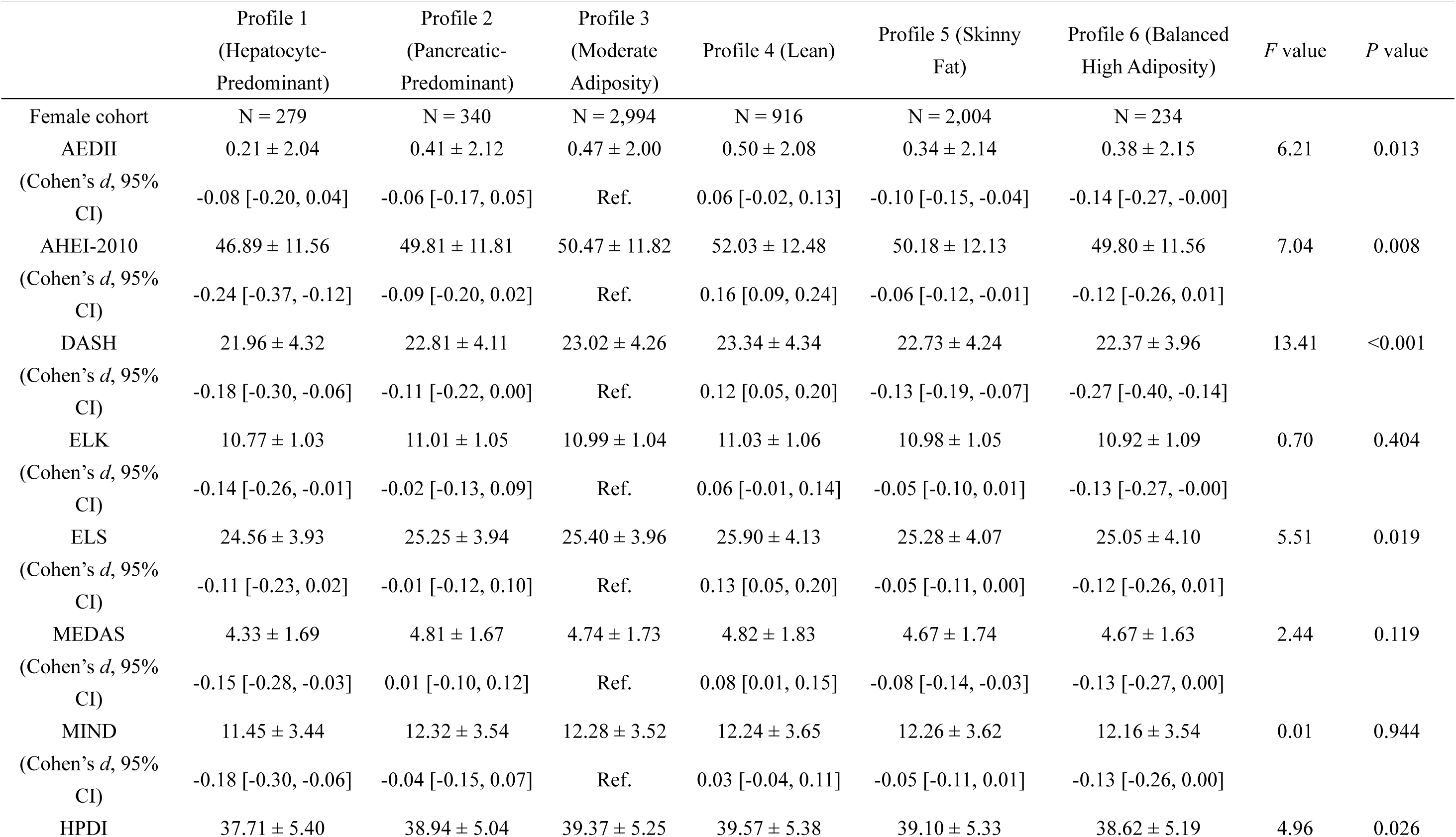
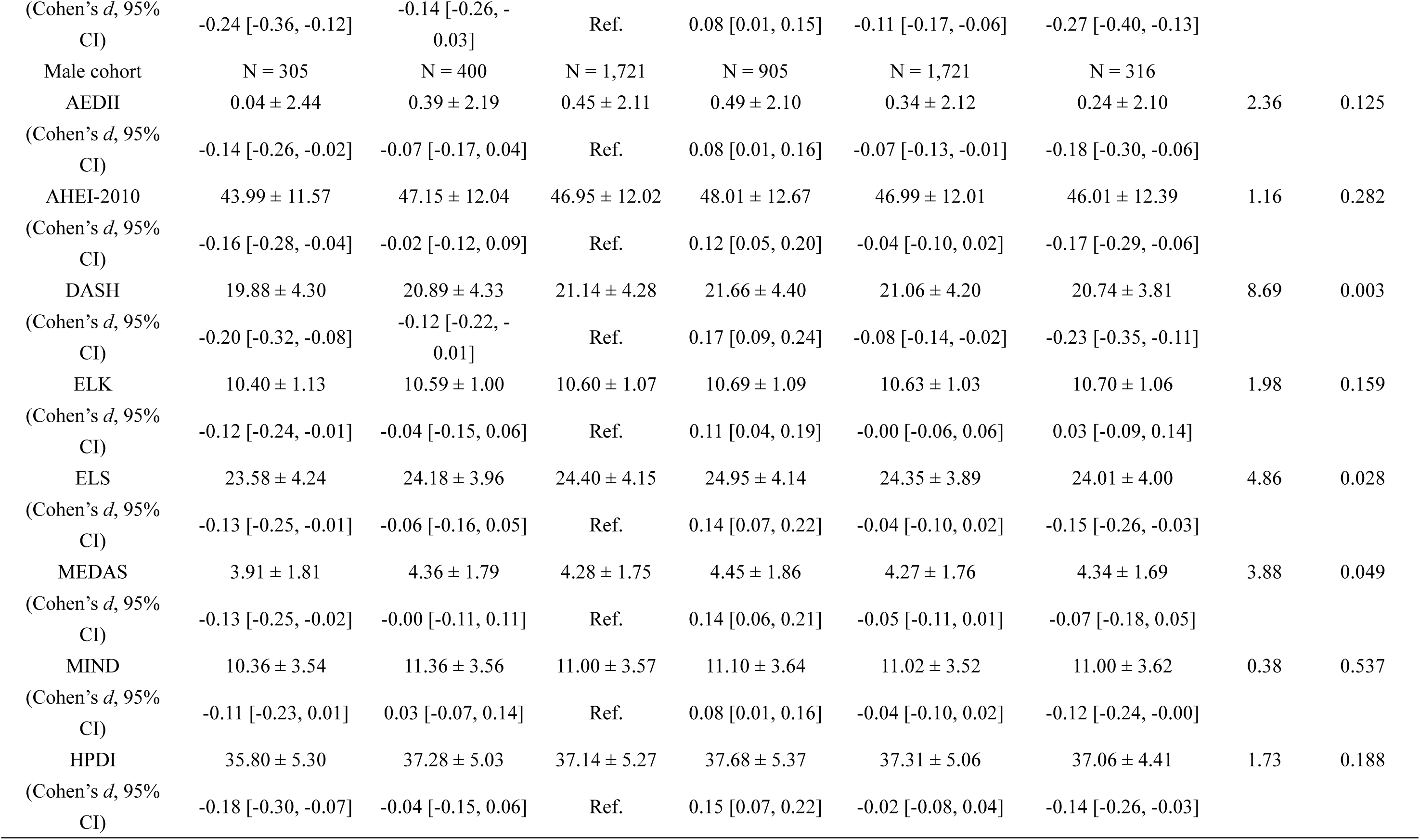
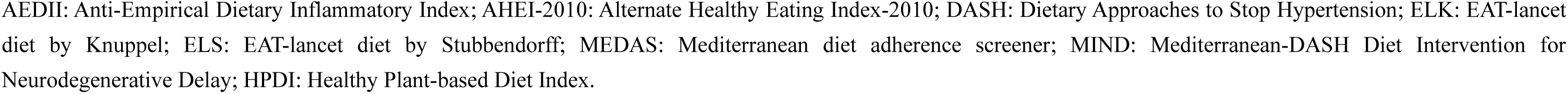
Sex-stratified comparisons of dietary pattern scores across latent profiles identified using MRI data.

### 3.7 The modulatory role of LPA profiles on the association between dietary pattern and myocardial infarction risk

To clarify whether the relationships between dietary patterns and clinical outcomes differed across distinct LPA-derived subtypes, we explored the modifying effects of LPA-derived subtypes on the associations between dietary scores and primary endpoints (**Fig. 5B**). Only dietary patterns had statistically significant relationships with the primary endpoints were included in the interaction analysis (marked with asterisks in **Fig. 3**). In the T2D cohort, a significant interactive association between the MIND diet score and Profile 6 was observed for myocardial infarction risk (FDR-adjusted *P* = 0.009). Similarly, in the CGD cohort, an interaction between the DASH diet score and Profile 6 was observed for myocardial infarction risk (FDR-adjusted *P* = 0.014). In summary, T2D patients are unlikely to benefit from the MIND diet, whereas CGD patients in Profile 6 derive no benefits from following the DASH dietary pattern.

### 3.8 Predictive performance of nomogram for primary endpoints

Time-dependent Cox regression models were developed to predict primary endpoints, incorporating demographic characteristics, dietary scores, and MRI-derived profiles. Following rigorous variable selection, final parsimonious models were transformed into quantitative nomograms, among which only the models for stroke, all-cause mortality, and myocardial infarction yielded statistically robust results **(Fig. 5C-5E**). Predictive performance was evaluated using time-dependent ROC (**Fig. 5F**), PRC curves (**Fig. 5G**), calibration curve (**Fig. S1A)** and DCA curve (**Fig. S1B)**.

For myocardial infarction prediction, the 3-year, 5-year, and 10-year risk models presented ROC-AUC values of 0.807, 0.788, and 0.766, alongside corresponding PRC-AUC values of 0.364, 0.355, and 0.357, respectively. For the stroke endpoint, the matched ROC-AUCs were 0.699, 0.706, and 0.708, with PRC-AUCs of 0.321, 0.353, and 0.357. Regarding all-cause mortality prediction, the 10-year and 15-year survival models achieved ROC-AUCs of 0.815 and 0.748, and PRC-AUCs of 0.376 and 0.341.

## 4. Discussion

This study aimed to investigate the prognosis of adverse clinical outcomes and optimize precision dietary strategies for patients with T2D, CGD, and their comorbidity. Significant positive correlation existed between T2D and CGD, and their comorbidity markedly raised risks of cardiovascular events, ESRD, dementia and all-cause mortality. The co-occurrence of T2D and CGD exerted synergistic effects on myocardial infarction and all-cause mortality, exacerbated systemic lipid metabolic disorders and multiorgan atrophy, while HPDI and DASH diets mitigated all-cause mortality risk by attenuating this synergistic interaction. Beneficial dietary patterns varied distinctly across T2D, CGD, and comorbid populations, with MEDAS, AEDII and DASH diets yielding optimal protective effects against corresponding adverse outcomes. Latent profile analysis identified a balanced high adiposity subtype that modified diet-related health associations, highlighting the value of phenotype-tailored dietary intervention. A combined nomogram integrating demographic, dietary and body composition data achieved reliable long-term predictive capability for myocardial infarction, stroke and all-cause mortality.

Based on a systematic search of PubMed, Embase, Web of Science, and the Cochrane Library, limited studies have specifically assessed the clinical prognosis of patients with concurrent T2D and CGD. We identified significant synergistic effects of T2D and CGD on myocardial infarction and all-cause mortality, highlighting the need for integrated dual-disease management rather than isolated care. The non-significant synergistic association for ESRD may be attributable to dominant T2D-related renal microvascular injury or insufficient event numbers (37), warranting cautious interpretation instead of definitively ruling out clinically meaningful effects. Collectively, our findings support integrating routine CGD screening and holistic care into long-term T2D management, alongside optimized glycemic control for CGD patients, to disrupt the T2D-CGD pathophysiological vicious cycle and reduce adverse clinical risks. Notably, high adherence to HPDI and DASH diets significantly attenuated the synergistic mortality risk of comorbid T2D and CGD, plausibly by suppressing chronic inflammation (38), ameliorating insulin resistance (39), and restoring intestinal mucosal integrity (40, 41). This study fills an important evidence gap by demonstrating modifiable dietary regulation of T2D-CGD disease interactions, providing mechanism-based precision nutrition insights to refine generic dietary guidelines for high-risk comorbid populations (42, 43).

Our findings confirm that optimal dietary strategies are subgroup-dependent. In patients with T2D only, the MEDAS diet confers superior benefits. Abundant in unsaturated fatty acids and polyphenolic compounds (44), it effectively ameliorates insulin resistance and systemic oxidative stress (45), the principal pathological features fueling T2D progression and its associated micro- and macrovascular complications (46, 47). For patients with isolated CGD, the AEDII diet exerts the strongest protective effects, as it directly targets chronic gastrointestinal mucosal inflammation, the central pathogenic mechanism of CGD (48). By suppressing local and systemic inflammatory cascades, this dietary pattern alleviates mucosal lesions and prevents subsequent adverse sequelae (38, 49, 50). In individuals with comorbid T2D and CGD, optimal dietary strategies vary according to clinical endpoints: AEDII for ESRD, ELS for dementia, and DASH for all-cause mortality. This heterogeneity may be explained by divergent molecular and physiological mechanisms underlying different dietary patterns, which ultimately translate into varied prognostic effects across distinct clinical outcomes.

This study expands on previous evidence (23) by integrating MRI-derived organ volumes along with fat distribution into the LPA framework, offering a more comprehensive characterization of metabolic body composition phenotypes. The Balanced High Adiposity phenotype was found to modify the effect of healthy diets on myocardial infarction risk. Patients with T2D in this phenotype were not significantly associated with benefit from the MIND diet, and similarly, those with CGD were not significantly associated with benefit from the DASH diet. This highlights the necessity of combined interventions aimed specifically at achieving substantial fat reduction. Abdominal MRI provides precise quantification of regional adiposity and organ volumes. However, its high cost, time requirements, and limited availability hinder immediate clinical use. Practical alternatives include anthropometric indices such as waist circumference, waist-to-hip ratio, and waist-to-height ratio. These measures are inexpensive, widely used, and correlate with visceral adiposity. Bioelectrical impedance analysis offers another accessible option (51). Modern multi-frequency segmental devices achieve high correlations with MRI-measured visceral adipose tissue, balancing reasonable accuracy with feasibility for large-scale screening and routine monitoring. While further validation is needed to confirm the utility of these proxies, clinicians may currently use anthropometric and bioelectrical impedance analysis measures to approximate MRI-derived body composition phenotypes.

Several limitations should be noted. First, the observational study design precludes causal inference, and residual confounding may persist despite extensive covariate adjustment. Thus, randomized controlled trials are warranted to confirm the prognostic benefits of dietary interventions in this comorbid population. Second, as the UK Biobank cohort is predominantly composed of White individuals with generally higher health awareness, the findings may not be generalizable to other ethnic or socioeconomic groups, necessitating validation in more diverse populations. Third, the MRI study had a limited sample size, particularly for the T2D-CGD subgroup. Therefore, the body composition phenotype findings require replication in larger imaging cohorts.

## Conclusion

Comorbid T2D and CGD synergistically increase the risk of myocardial infarction and all-cause mortality, and healthy diets were associated with reduced all-cause mortality risk by attenuating this synergistic interaction. Given that optimal dietary approaches differ across populations and MRI-identified LPA phenotypes, targeted nutritional interventions tailored to specific phenotypes are recommended.

## Author Contributions and Guarantor Statement

B.H., Y.Y.L., and H.X. were involved in the conception, design, and conduct of the study and the analysis and interpretation of the results. B.H. wrote the first draft of the manuscript, and all authors edited, reviewed, and approved the final version of the manuscript. B.G., G.B.C, and Y.Y. are the guarantors of this work and, as such, had full access to all the data in the study and take responsibility for the integrity of the data and the accuracy of the data analysis. X.W.Y., A.L.Y., Y.X.J., and S.R.L. provided clinical expertise and validation. L.J.D., S.G., Y.T., and X.Y.B. contributed to data verification and visualization. M.H.N. and L.F.Y. performed validation analysis.

## Data Availability Statement

All data are available from the UK Biobank (https://www.ukbiobank.ac.uk/).

